# Cognitive and Neuroimaging for Neurodegenerative Disorders: A Cohort Study Design with Initial Findings

**DOI:** 10.1101/2025.05.19.25327923

**Authors:** Akram A. Hosseini, Beili Shao, Abigail Rebecca Lee, Permesh Dhillon, Kehinde Junaid, Bruno Gran, Peter Sellars, Hannah Sargisson, JeYoung Jung, Elizabeta Mukaetova-Ladinska

**Author notes:** **Correspondence:** Dr Akram A. Hosseini.

## Abstract

**Introduction:** Dementia presents with significant heterogeneity across age groups, particularly in early-onset cognitive decline (EOCD), which poses diagnostic and management challenges. The Cognitive and Neuroimaging for Neurodegenerative Disorders (CogNID) study aims to characterise clinical, cognitive, neuroimaging, and biomarker features across a diverse cohort of individuals with cognitive impairment, with a focus on diagnostic complexity, biomarker utility, and mortality. **Methods:** Out of 680 study participants within this prospective cohort enrolled from the real-world clinics within the National Health Service, who consented to take part in the study, we analysed data from 429 individuals recruited between December 2018 and November 2024 from the Memory Clinics, including the young-onset dementia service and associated services. Participants underwent structured cognitive assessments, neuroimaging (MRI/CT), and Cerebrospinal fluid (CSF) biomarker evaluation, where available. Diagnoses were made by multidisciplinary consensus. Group comparisons were conducted between early-onset (EOCD, <65 years) and late-onset cognitive decline (LOCD, ≥65 years). **Results:** Of the 429 participants, 349 (81.4%) had EOCD and 80 (18.6%) had LOCD. The mean age was 60.05 years, with no significant difference in sex or ethnicity across groups. Depression and anxiety were common (29.6%), as were cardiovascular risk factors. Lumbar punctures were more frequently performed in EOCD (p = 0.03), with 36.4% of tested participants demonstrating biomarker profiles consistent with Alzheimer’s disease (A+T+). Functional cognitive disorder (FCD) was more common in EOCD (22.3% vs. 5.0%, p < 0.001). Subgroup analysis revealed significantly lower ACE-III scores and higher pathological CSF findings in Alzheimer’s disease versus FCD. Mortality was higher in the LOCD group (11.3% vs. 4.6%, p = 0.03). **Conclusion:** The CogNID study highlights the clinical and diagnostic heterogeneity of individuals with cognitive impairment, particularly in younger adults. Incorporating neuroimaging and CSF biomarkers into routine clinical pathways enhances diagnostic precision and reveals distinct phenotypic profiles between EOCD and LOCD. These findings underscore the need for harmonised diagnostic protocols, broader biomarker accessibility, and inclusive recruitment strategies in dementia research and clinical services.

## Introduction

### Background

Early-onset Cognitive Decline (EOCD) poses a distinct clinical challenge, especially due to the diversity in underlying causes and the complexity in making an accurate diagnosis. Although it is estimated that 70,800 people in the UK have been diagnosed with early-onset dementia (Dementia UK, 2022), this number is likely to be a significant underestimate. The true figure could be much higher, with a potential 19,000 undiagnosed cases in England alone (Alzheimer’s Society, 2023). Dementia in adults under the age of 65 years can present differently to late-onset dementia, particularly regarding psychiatric and behavioural symptoms (Tsoukra et al., 2021). This can lead to misdiagnosis, or a delay in reaching a EOCD diagnosis. A review of the 2019 English national memory service audit found that those under the age of 65 years were around 40% less likely to get a correct dementia diagnosis (Cook et al., 2021). Compounding the difficulty, EOCD accounts for just 119 per 100,000 cases (Hendriks et al., 2021) compared to functional disorders like schizophrenia, which presents at 1 per 100 cases (Rethink Mental Illness, 2024).

In younger populations, the aetiological diversity is greater compared to older adults, where Alzheimer’s disease (AD) accounts for most dementia cases. In younger adults, AD represents less than half of diagnoses, with frontotemporal lobar degeneration (FTLD) and other rarer disorders contributing to over 30% of cases (Harvey et al., 2003). In a review from Hendriks, the prevalence of AD in adults aged 30–64 years was estimated as 54.1 per 100,000 in the European population. Vascular dementia was estimated as 19.5 per 100,000 in the European population and FTLD was 2.9 per 100,000 (Hendriks et al., 2021).

EOCD has a profound impact not only on the individual but also on their family and wider social network. Individuals are often amid their working lives, and many face early job loss or forced retirement, leading to financial hardship and a loss of personal identity. Unlike those with late-onset dementia, individuals with EOCD may have dependent children or caregiving responsibilities, which places additional emotional and logistical burdens on families (van Vliet et al., 2010). These challenges highlight the urgency of early and accurate diagnosis, access to appropriate services, and the development of preventative strategies to reduce the wider impact of the disease (Spreadbury et al., 2018).

There is a growing evidence base to support the role of modifiable risk factors in the development dementia. In the Lancet Commission on dementia prevention, intervention and care, they estimated that 45% of dementia cases may be preventable by addressing the risk factors throughout the life span, such as smoking, alcohol consumption, and education level (Livingston et al., 2024). This complexity, along with emerging concerns such as cognitive impairment related to COVID-19, highlights the need for robust, multifaceted diagnostic approaches.

Longitudinal studies provide fundamental information in understanding the diverse population living with dementia, and their diagnosis and care journey. These studies enable researchers to explore the biological, social, and environmental factors which affect individuals and better understand how these interact to affect cognitive decline (Khalatbari-Soltani et al., 2024). However, longitudinal data often collected through cohort studies is not readily available, and research has been limited in certain parts of the globe (Khlatbari-Soltani et al., 2024).

The UK National Health System represents a valuable source of clinical data, including longitudinal data from real-world clinical practice. Incorporating research models with ethically approved protocols and consents compliant with the General Data Protection Regulation (GDPR) would enable harmonised data collection and effective use of this resource. Considering recent advanced and multi-modal imaging and biomarker investigations, as well as emerging therapies, integrating research frameworks and ethically approved consent models into NHS data collection would be crucial to support the implementation of clinical guidelines, reduce healthcare burden, and centralise data collection. This would enable higher-impact research, enhance training, and ensure clinical studies are grounded in robust, meaningful real-world data.

### Neuropsychological Assessment

Neuropsychological assessments are used routinely to provide objective evidence of cognitive difficulties (O’Malley et al., 2019). However, existing neuropsychological tools are often insufficient in the early and accurate diagnosis of dementia, especially in patients with EOCD (O’Malley et al., 2019). The Addenbrooke’s Cognitive Examination III (ACE-III; Hsieh et al., 2013) is often used in clinical settings to detect any change in cognition. The ACE-III has a maximum score of 100 and questions explore five cognitive domains; memory, attention, language, verbal fluency, and visuospatial function (Hsieh et al., 2013). Evidence suggests that the ACE-III is more sensitive that other assessments, such as the Mini Mental State Examination, in the detection of dementia (Bruno et al., 2019). Although the ACE-III has become routinely used in the UK memory clinics, it is important to consider demographic variables and modifiable risk factors for dementia when interpreting scores (Bruno et al., 2019).

While neuropsychological assessments are helpful in identifying any initial cognitive changes, their scores should not be considered alone when diagnosing dementia (O’Malley et al., 2019). Clinical assessments in isolation are inaccurate in 30-35% of cases (Beach et al., 2012). The incorporation of biomarkers and neuroimaging is essential for establishing a multidimensional diagnostic approach, improving the likelihood of accurate diagnosis and reducing the risk of misdiagnosis or underdiagnosis, particularly for patients with EOCD (O’Malley et al., 2019).

### Rationale

#### The aim of this study was twofold

First, to standardise cognitive assessments, neuroimaging, and biomarker investigations for patients referred to neurology-led tertiary referral clinics. This initiative was designed to support regional memory clinics by implementing a harmonised protocol for cognitive evaluation, neuroimaging, and the use of biomarkers in accordance with NICE guidelines. This approach was particularly important during a period of clinical uncertainty regarding the indications or lack of a nationwide accessibility for Cerebral Spinal Fluid (CSF) biomarkers or Amyloid-PET imaging in routine UK clinical practice.

Secondly, to establish a research platform that would maximise the use of underutilised but costly investigations performed through the National Health Service. This platform aimed to generate real-world clinical data to inform demographic profiles, particularly of populations commonly under-represented in research. It also sought to leverage clinical and neuroimaging data for ongoing and future studies. In addition, individuals with mild cognitive impairment (MCI) were offered opportunities to participate in parallel and future research. For Participants undergoing diagnostic lumbar puncture, the option to donate additional CSF samples for research was provided, enabling complementary studies on neurodegenerative biomarkers.

In this article, we introduce the Cognitive and Neuroimaging for Neurodegenerative Disorders (CogNID) Study, a longitudinal observational clinical study involving participants experiencing cognitive difficulties. We present the outline of the study, the measures we use, and the initial findings from the study’s six years of recruitment and data collection.

### Objectives

#### Primary Objectives

1. To identify cognitive and neuroimaging markers that can assist in the early diagnosis of neurodegenerative disorders. This includes assessing predictors of disease progression and identifying risk factors within diverse ethnic communities and across the lifespan, with a particular focus on younger patients.
2. To establish a diverse research platform that includes an EOCD alongside a control group, enabling further research into neurodegenerative disorders and providing a foundation for longitudinal studies that integrate clinical, imaging, and biomarker data.

### Secondary Objectives

1. To evaluate the relationship between clinical features and neuroimaging findings, aiming to identify patterns that correlate with early markers of neurodegenerative disorders.
2. To track the progression of cognitive decline and corresponding neuroimaging changes over time, facilitating the identification of biomarkers associated with disease trajectory.
3. To identify risk factors for neurodegenerative conditions by incorporating a control group, allowing comparative analysis and examination of factors that may modify disease progression.
4. To share data with open-access repository datasets, integrating cognitive, neuroimaging, and biomarker data to enable broader research collaboration and facilitate secondary analyses

## Materials and Methods

### Ethics approval

This study has been reviewed and approved by the NHS East Midlands Derby Research Ethics Committee (18/EM/0292; IRAS: 250525). All amendments and protocol changes have been reviewed and authorised by Nottingham University Hospitals NHS Trust and the above ethics committee, before being implemented. The study has also been reviewed and approved by the Research & Innovation Department of Nottingham University Hospitals NHS Trust. The CogNID Study has been accepted by ClinicalTrials.gov (NCT03861884). All participants must provide written informed consent prior to any data collection.

### Study Design and Setting

Research activities for this cross-sectional and longitudinal observational cohort study were carried out by researchers and clinicians working in the Queen’s Medical Centre, Nottingham University Hospitals NHS Trust, and the University of Nottingham. All cognitive assessments were carried out in-person within Nottingham University Hospitals NHS Trust, and conducted by either a clinician, assistant psychologist, or trained research practitioners.

This study initially focused on recruiting EOCD who were primarily referred from primary or secondary care within the region. However, as often occurs in real-world clinical practice, patients from older age groups, cognitive consequences related to COVID-19, and those referred from other regions were also included. In response, the study protocol was modified. Consequently, 80 late-onset dementia cases (18.65%) were incorporated into this cohort.

## Recruitment

Participants were recruited from memory clinics based at Nottingham University Hospitals NHS Trust, Nottinghamshire Healthcare Foundation Trust, Leicester Young Age Dementia Services. An Additional older age control group were recruited from Cognitive Disorders clinics Nottingham University Hospitals NHS Trust. Patients attending these clinics have been referred by primary and secondary care services within the East Midlands region including Nottinghamshire, Derbyshire, Leicestershire as well as Lincolnshire. Nottingham University Hospitals NHS Trust is also a tertiary referral centre for the cerebrospinal fluid (CSF) diagnostic process, and some participants were recruited via this pathway.

Eligible and interested patients received a Participant Information Sheet outlining the background and purpose of the study to read and were given the opportunity to ask questions and then complete and sign an Informed Consent Form. Some participants were co-recruited for a study involving blood tests and CSF examinations (MREC reference 08/H0408/167). Additional blood and CSF samples were donated for research purposes.

## Inclusion and Exclusion Criteria

All potential participants need to have the ability and capacity to give consent prior to study enrolment. Capacity was based on the criteria defined by the Mental Capacity Act 2005, and any queries pertaining to an individual’s capacity were resolved through discussions with a medical professional.

### Inclusion Criteria

- Aged ≥18 years and ≤ 75 years. We modified it to Aged >18 to include older age for all eligible participants last year.
- Experiencing cognitive symptoms or cognitive impairment and referred for clinical assessments due to this;
- Pathological, genetic, or imaging biomarker evidence to suggest FTLD, AD or Vascular Cognitive Impairment (VCI); **OR**
- A clinical diagnosis of one of the following which fall under the named conditions above:
  - FTLD – Frontotemporal dementia; behavioural variant, semantic dementia, progressive non-fluent aphasia, progressive apraxia of speech; Progressive supranuclear palsy; Corticobasal degeneration; Poorly differentiated tauopathy with compound features of FTLD;
  - AD – Amnestic (classic) Alzheimer’s disease; Posterior cortical atrophy; Logopenic or mixed primary progressive aphasia; Frontal variant Alzheimer’s disease, and Limbic-predominant Age-related TDP-43 Encephalopathy;
  - Dementia with Lewy Bodies (DLB) and Parkinson’s Disease Dementia (PDD);
  - VCI – Hereditary vascular dementia; multi-infarct dementia; post-stroke dementia; subcortical ischaemic vascular disease; cerebral amyloid angiopathy spectrum; mild cognitive impairment;
  - MCI secondary to any neurological or metabolic disorder; suspected neurodegeneration with brain iron accumulation; NBIA (genetic neurological disorders characterised by abnormal accumulation of iron in the basal ganglia)

### Exclusion Criteria

- Unable to provide informed consent;
- Not speaking English before the age of five years; This criterion was removed to include any spoken language and ensure inclusivity of ethnic minorities last year.
- Presence of conditions such as learning disability, substance misuse, major depression, schizophrenia, or use of medications known to significantly impact the study participant’s cognition;
- Unable to sustain concentration for at least 30 minutes, in single sitting.

### Assessments

Comprehensive data collected across multiple domains for participants enrolled in the study (Figure 1). Once consented, all participants underwent an ACE-III cognitive assessment during, or shortly after, their first visit to a clinic. If a participant has recently completed an ACE-III with an experienced clinical team member, a copy of this assessment was used instead. There were alternative cognitive assessments available within the clinic, if required. These included the Montreal Cognitive Assessment (MoCA; Nasreddine et al., 2005); the Birmingham Cognitive Screen (BCoS; Humphreys et al., 2012); and the Uniform Data Set (UDS; Besser et al., 2018). If low mood was identified during the clinic visit, it was assessed with the Hospital Anxiety and Depression Scale (HADS; Zigmond et al., 1983). The Neuropsychiatric Inventory (NPI; Cummings et al., 1994) was used as an optional assessment, should the clinician wish to further explore the psychopathology of symptoms.

**Figure 1.**
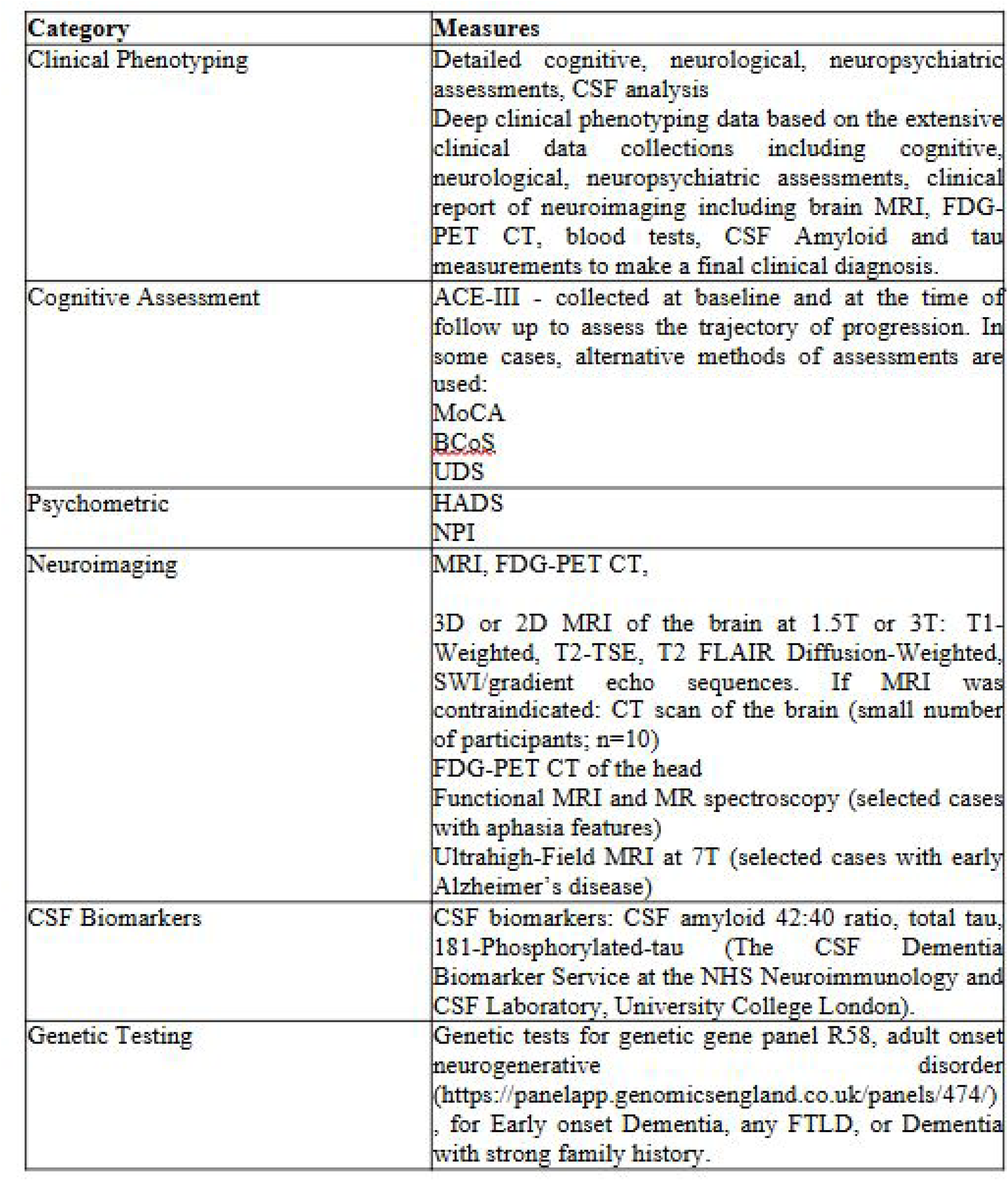
Overview of Clinical, Cognitive, Neuroímaging, and Biomaiker Measures in the Study Cohort. Abbreviations: ACE-EIĪ = Addenbrooke’s Cognitive Examination-III: MoCA = Montreal Cognitive Assessment; BCoS = Birmingham Cognitive Screen; UDS = Uniform Data Set; HADS = Hospital Anxiety and Depression Scale; NPI = Neuropsychiatric Inventory; CSF = cerebrospinal fluid: FDG-PET = fluorodeoxyglucose positron emission tomography: MRI = magnetic resonance imaging: CT = computed tomography; SWI = susceptibility-weighted imaging: DWT = diffusion-weighted imaging; FTLD= frontotemporal lobar degeneration.

Participants consented to share their clinical data, which includes pseudonymised images and clinical report of 1.5T or 3T MRI, 7T MRI, CT brain scans, FDG-PETCT brain scans, the results of diagnostic lumbar punctures, and biochemical blood tests. These tests encompass cell count, renal and liver function tests, blood glucose, thyroid function, vitamin B12, and folate levels, as well as, in selected cases, autoimmune encephalitis antibodies and infection screens.

Imaging protocol for the brain MRI included 3D structural T1-Weighted, T2-Weighted, FLAIR, Diffusion-weighted, and gradient echo sequences at 3T or 1.5T MRI. Participants may consent to undergo advanced neuroimaging modalities, such as MR spectroscopy, functional MRI or ultra-high-field MRI at 7T (Hari et al., 2024; Adeyemi et al., 2024). Additional advanced neuroimaging was undertaken within Nottingham University Hospitals NHS Trust or the Sir Peter Mansfield Imaging Centre, University of Nottingham.

Diagnostic lumbar puncture provides information on CSF biomarkers, including the Amyloid β (Aβ) 1-42:40 ratio, total tau, and 181-phosphorylated tau (Hosseini et al., 2022). However, in the initial stages of the study, the national laboratory testing for CSF biomarkers only reported Aβ 1-42 without Aβ 1-40. As a result, this Aβ 1-42:1-40 ratio wss unavailable for selected cases.

### Follow-up Assessments

Participants were followed up at 6 and 12 months, then annually, with optional repeat imaging and psychometric assessments. Follow-ups may be conducted in-person, via telephone, or remotely. Repeat MRI scans arranged one year after recruitment for participants who consent.

## Statistical Methods

Descriptive statistics were used to summarise demographic, cognitive, and biomarker data. Continuous variables were assessed for normality and were presented as means with standard deviations (SD) for normally distributed data, or as medians with interquartile ranges (IQR) for non-normally distributed data, as appropriate. Statistical analyses were conducted to compare demographic and clinical characteristics between the early-onset and late-onset dementia groups. Independent samples t-tests were used for continuous variables that met the assumption of normality, while the Mann–Whitney U test was applied to non-normally distributed data. Categorical variables were assessed using chi-square tests; where expected cell counts were less than five, Fisher’s exact tests were employed. A p-value of less than 0.05 was considered statistically significant. All analyses were performed using Python, with the scipy.stats libraries.

## Results

### Recruitment and Demographics

Between December 2018 and April 2025, a total of 680 participants, who attended our memory clinics, consented to take part in the clinical study for research in dementia. Of these, the data until November 2024 were analysed in this report. After excluding 19 healthy controls, the final analytical cohort comprised 429 individuals with a clinical diagnosis of dementia (see Figure 2). The mean age of participants was 60.05 years (range 18–84), with a slight male predominance (54.6%). The majority were of Caucasian ethnicity (63.9%). Regarding lifestyle factors, 5.1% of participants were current smokers, 26.3% reported current alcohol consumption, and 2.3% reported excessive alcohol use (see Figure 3).

**Figure 2.**
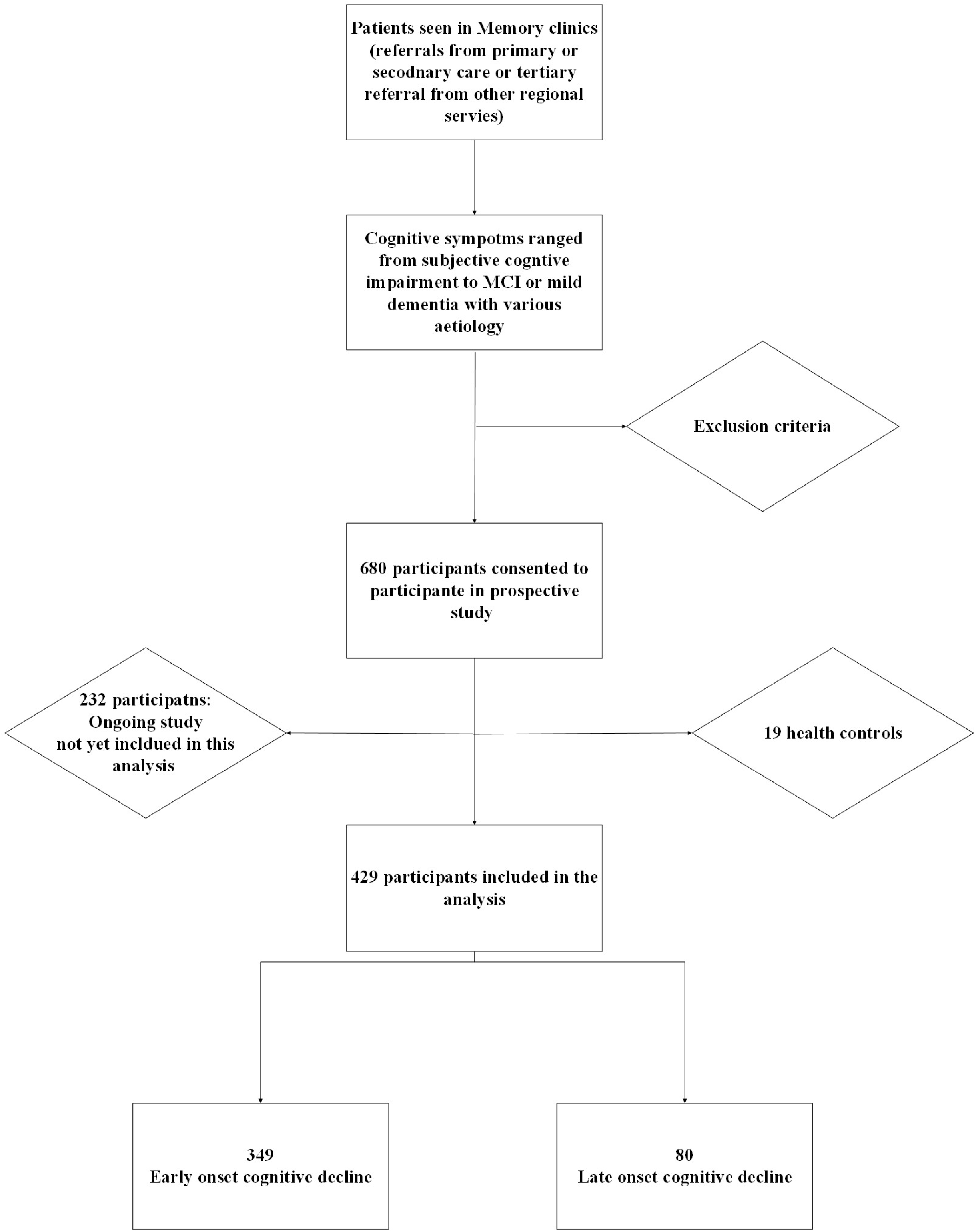
Recruitment Flowchart.

**Figure 3.**
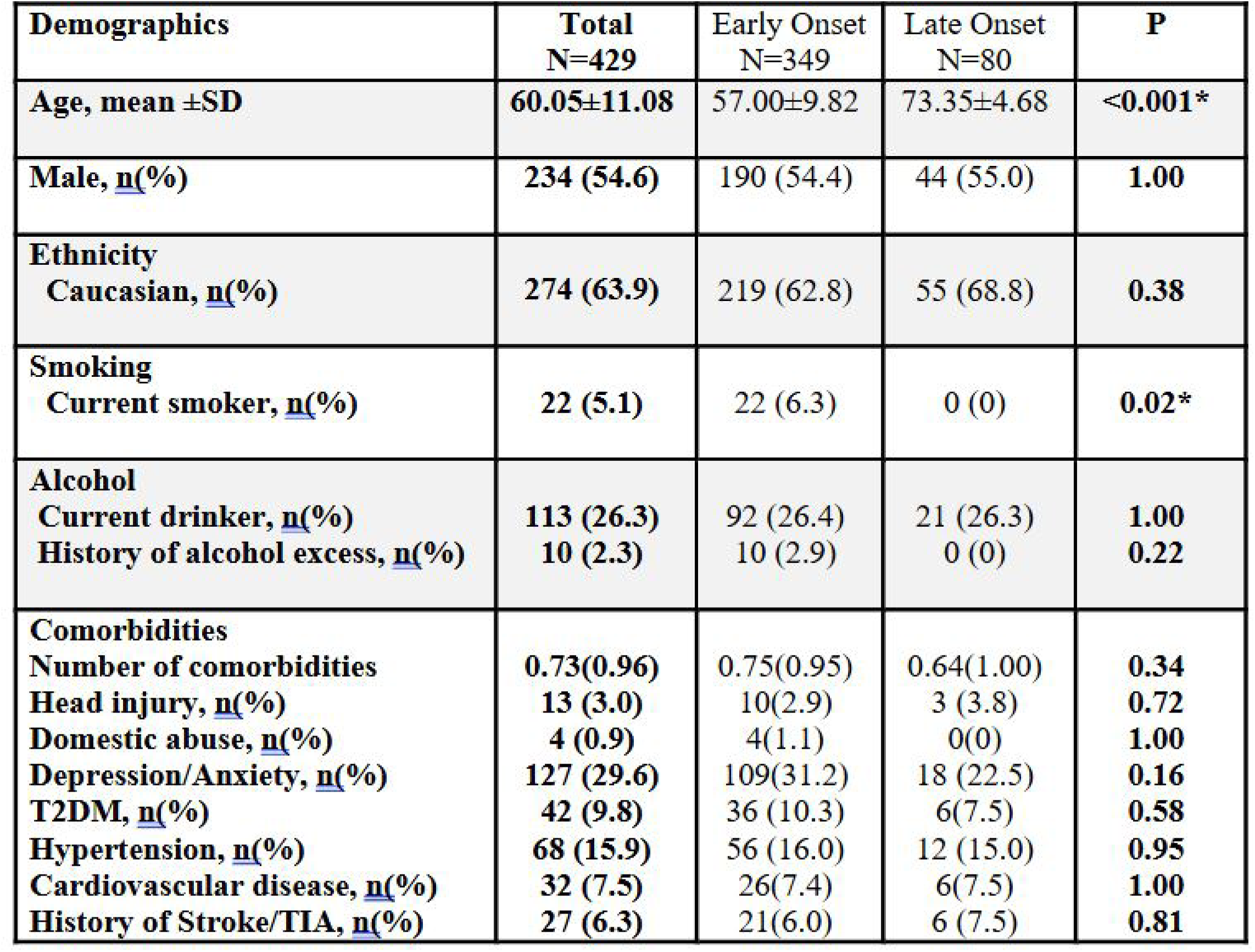
Demographic and clinical characteristics of participants with early-onset and late-onset cognitive decline. Demographic and clinical variables are summarized for the full cohort (N=429). early-onset dementia group (N=349), and late-onset dementia group (N=80). Continuous variables are presented as mean ± standard deviation (SD); categorical variables are presented as counts with percentages. Age is shown as mean ± SD with range. P-values represent comparisons between early-onset and late-onset groups using independent samples t-tests for continuous variables, and Chi-square or Fisher’s exact tests for categorical variables, as appropriate. Asterisks (*) indicate statistically significant differences at *p* < 0.05. T2DM - Type 2 diabetes mellitus; TIA - Transient ischaemic attack.

Out of the recruited 429 individuals, 349 (81.4%) had EOCD and 80 (18.6%) had LOCD. The mean age in the EOCD was 57.00 ± 9.82 years, whereas the LOCD was 73.35 ± 4.68 years (p < 0.001). The proportion of males was similar between groups (54.4% early-onset vs. 55% late-onset; p = 1.0). Caucasian ethnicity was the most common across both groups, with no significant difference between EOCD (62.8%) and LOCD (68.8%; p = 0.38). Current smoking was significantly more prevalent in the EOCD group (6.3%) compared to none in the LOCD group (p = 0.02). There were no significant differences in alcohol consumption or history of alcohol excess between groups.

The demographic data revealed notable patterns of comorbidities among the 429 individuals in the cohort, emphasizing the multifactorial health challenges present in the population. Depression and anxiety were the most prevalent comorbid conditions, affecting 127 individuals (29.6%), which aligns with their frequent association with chronic and neurological conditions. Hypertension, a common risk factor for cardiovascular and neurological diseases, was present in 68 individuals (15.9%), while Type 2 Diabetes Mellitus (T2DM) affected 42 individuals (9.8%). Cardiovascular disease was reported in 32 individuals (7.5%), reflecting a significant burden of systemic health issues. Additionally, 27 individuals (6.3%) had a history of stroke or TIA, indicating a notable prevalence of cerebrovascular complications. Less common comorbidities included head injury (13 individuals, 3.0%) and a history of domestic abuse (4 individuals, 0.9%). These findings highlight the importance of addressing mental health, metabolic disorders, and cardiovascular risks in managing this population comprehensively (Livingston et al., 2024).

Co-existing medical and mental health conditions (i.e. rates of depression/anxiety, T2DM, hypertension, cardiovascular disease, and history of stroke/TIA) were comparable between EOCD and LOCD groups, with no statistically significant differences observed (Figure 3).

### Neurocognitive Assessment, Neuroimage and Biomarkers

The median total ACE-III score among all participants was 73 (Figure 4). A threshold score of <82 was used to identify possible MCI, as suggested by Hsieh et al. (2013), and was observed in 72.2% of the cohort. The median sub-scores were as follows: Attention – 14, Memory – 15, Fluency – 8, Language – 23, and Visuospatial skills – 14. Age-specific cut-offs will be considered in subsequent analyses, in line with recommendations by previous study (Matías-Guiu et al., 2014).

**Figure 4.**
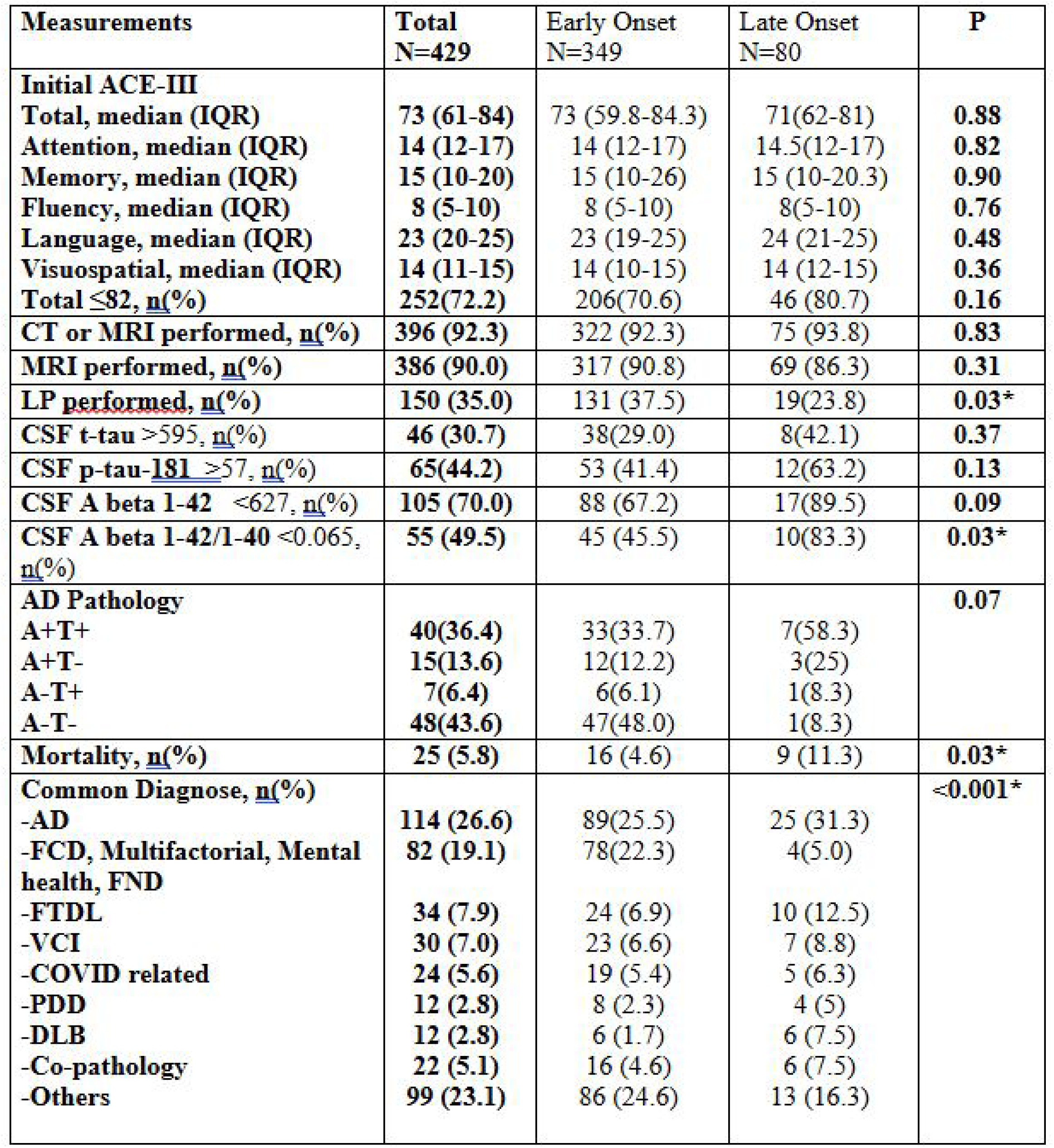
Details of Cognitive assessments, neuroimaging investigations and biomarkers among different neurodegenerative disorders within the study population. Abbreviations: ACE-III - Addenbrooke’s Cognitive Examination - III: CSF - cerebrospinal fluid: Aβ = amyloid-beta: t-tau = total tau: p-tau = phosphorylated tan: AD = Alzheimer’s disease: FTLD = frontotemporal lobar degeneration: VCI = vascular cognitive impairment: DLB = dementia with Lewy bodies; FCD = functional cognitive disorder: FND - functional neurological disorder. PPD = Parkinson’s Disease Dementia: DLB = Dementia with Lewy Bodies. CSF biomarker units: Aβl-42, t-tau, and p-tau-181 concentrations are reported in pg/mL, Aβ1—42/1-40 is a unitless ratio.

92.3% participants had at least one form of neuroimages performed including CT or MRI head. MRI head scans were performed in 386 participants (90.0%). CSF analysis was completed in 150 participants (35.0%). Among those with available CSF data, elevated total tau (t-tau >595 pg/ml) was present in 30.7%, elevated phosphorylated tau (p181-tau >57 pg/ml) in 44.2%, reduced Aβ1-42 (<627 pg/ml) in 70.0%, and a reduced Aβ1-42/1-40 ratio (<0.065) in 49.5%. An additional 91 participants with available CSF samples were recruited during the manuscript preparation phase.

Biomarker confirmed AD was defined as presents with amyloidosis and Tauopathy evident with reduced Aβ1-42/1-40 and increased p-181-tau protein in CSF (Jack et al., 2018). 135 participants had diagnostic lumbar puncture performed. However, Aβ1-40 was not tested in cases initially due to test accessibility. Out of 110 cases had both p-tau-181 and Aβ1-42/1-40, 40 cases (36.4%) fulfil these criteria with A+T+. 15 cases had amyloidosis without tauopathy, while 7 cases had tauopathy without amyloidosis.

Cognitive performance, as measured by ACE-III total and subdomain scores, did not significantly differ between EOCD and LOCD groups. MRI was performed frequently and comparably across both groups. However, lumbar punctures were significantly more common in the EOCD (p = 0.03), potentially reflecting greater diagnostic complexity or a wider differential diagnosis in younger individuals. CSF biomarker profiles revealed broadly similar levels of t-tau and p-tau-181 between groups. Although a higher proportion of LOCD participants exhibited low Aβ1-42 levels, this did not reach statistical significance (p = 0.09). In contrast, the proportion of participants with a pathological Aβ1-42/1-40 ratio (<0.065) was significantly greater in the LOCD group, suggesting a higher burden of Alzheimer’s pathology in older individuals.

### Diagnoses

The wide spectrum of diagnoses has been observed in this study, suggesting the complexity of differential diagnosis in dementia service (Figure 4). Diagnosis distribution was significantly different between EOCD and LOCD groups (p<0.001). AD was the most prevalent diagnosis across the cohort (26.6%), with a higher frequency in the late-onset group (31.3%) compared to early-onset cases (25.5%). Functional cognitive disorder (FCD) and psychiatric causes, encompassing multifactorial, psychiatric, and functional presentations, were the second most common overall (19.1%) and significantly more frequent in early-onset cases (22.3%) than in late-onset (5.0%). FTLD, VCI, and COVID-related cognitive syndromes were present across both groups with similar distribution. PDD and DLB were more commonly diagnosed in the late-onset group (6.3% and 7.5%, respectively), relative to early-onset (2.3% and 1.7%). Co-pathology was also more prevalent among late-onset cases, while other diagnoses were observed more frequently in EOCD group. The two most common combinations were AD + FTLD (n=7) and AD+DLB (n=3). Overall, there was a statistically significant difference in diagnostic distribution between early- and late-onset dementia groups (p < 0.001), reflecting distinct clinical profiles based on age of onset. Mortality was higher in the LOCD group (11.3% vs. 4.6% in EOCD, p=0.03), suggesting a poorer prognosis despite similar cognitive impairment severity and comorbidity burden.

### Alzheimer’s Disease and Functional Cognitive Decline

In this sub-analysis comparing patients diagnosed with AD (N=114) and FCD (N=55), several key demographic and clinical differences were found (Figure 5). Patients with FCD were significantly younger than those with AD (mean age 58.44 vs. 63.27 years; p < 0.001). There were no significant differences in sex distribution, ethnicity, smoking status, or alcohol consumption between groups. MRI brain was performed at similar rates, as was CSF biomarker testing. There was no mortality in the FCD group, compared with a 6.1% mortality rate in the AD group).

**Figure 5.**
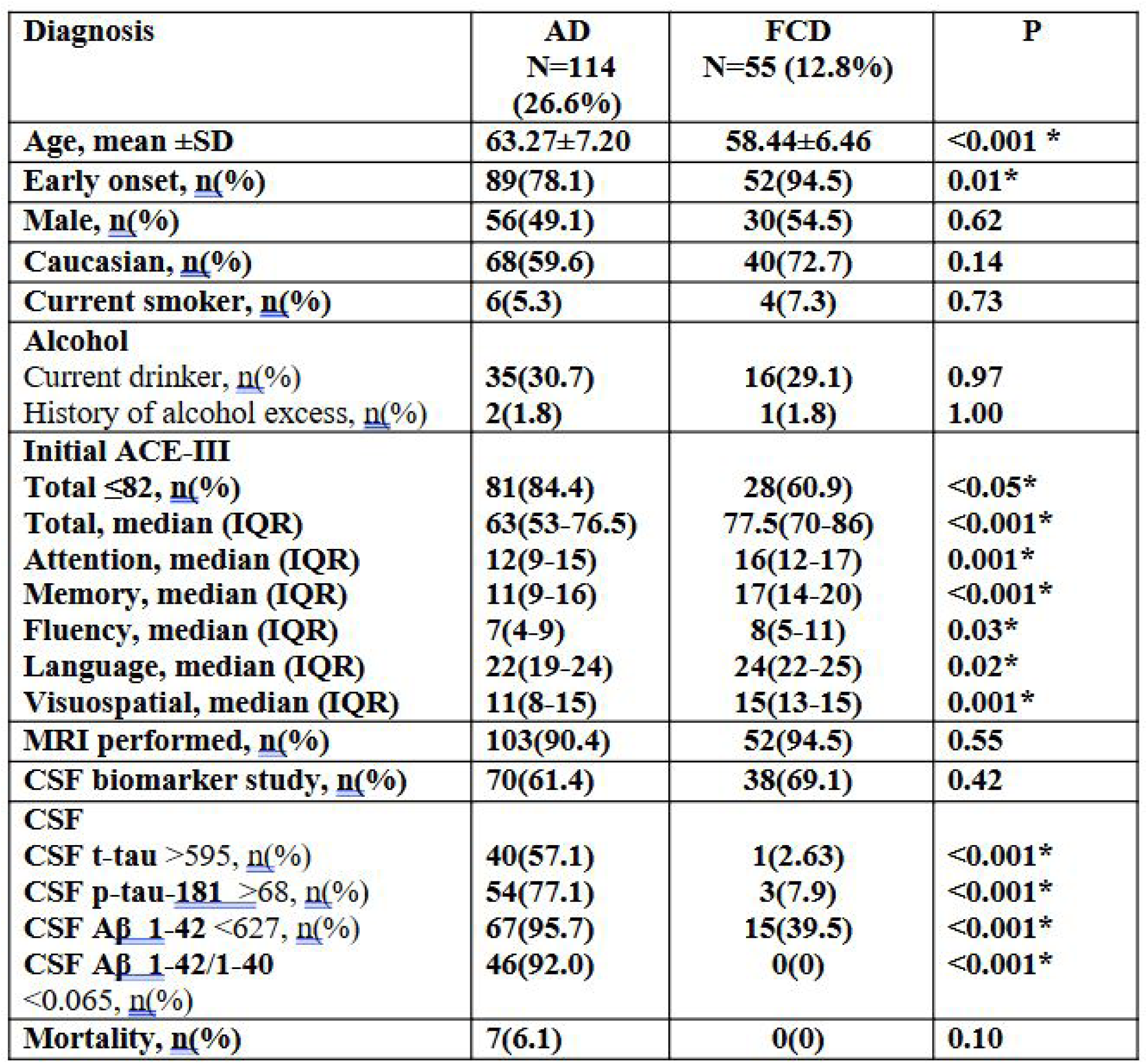
Demographic, cognitive, and CSF profiles of the most two common diagnoses among the study population. Abbreviations: ACE-IĪI = Addenbrooke’s Cognitive Examination - III; CSF = cerebrospinal fluid: Aβ = amyloid-beta; t-tau = total tau; p-tau = phosphorylated tau: AD = Alzheimer’s disease; FCD = functional cognitive disorder. CSF biomarker units: Aβl-42, t-tau, and p-taul8l concentrations are reported in pg/mL Aβ1–42/1–40 is a unitless ratio.

At initial assessment, participants with AD showed significantly lower cognitive performance across all domains of the ACE-III compared to those with FCD. A higher proportion of AD scored below the diagnostic threshold of 82 (84.4% vs. 60.9%, *p* < 0.05). The median total ACE-III score was notably lower in AD (63 [IQR 53–76.5]) than in the FCD group (77.5 [IQR 70–86], *p* < 0.001). Domain-specific analysis revealed significantly reduced scores in attention, memory, and visuospatial functions (all *p* ≤ 0.001), with additional but more modest differences in fluency (*p* = 0.03) and language (*p* = 0.02). These results suggest a broader and more severe cognitive profile at presentation among individuals with AD.

CSF biomarker profiles showed marked differences between AD and FCD participants. Abnormal levels of total tau (>595 pg/ml) were observed in 57.1% of AD participants, compared to only 2.6% in the FCD group (*p* < 0.001). Similarly, elevated phosphorylated tau (p-tau-181 >57 pg/ml) was present in 77.1% of AD cases versus 7.9% in the FCD group (*p* < 0.001). Reduced Aβ1-42 (<627 pg/ml) levels, indicative of amyloid pathology, were detected in 95.7% of AD cases, while only 39.5% of FCD participants showed this abnormality (*p* < 0.001). Additionally, 92.0% of AD participants had a reduced Aβ1-42/1-40 ratio (<0.065), compared to none in the FCD group (*p* < 0.001).

### Mortality

Among the 25 participants who died during the study period, the mean age at death was 65.28±11.22 years, with the majority being male (72%) and 48% identifying as Caucasian (Figure 6). The mean duration of follow-up prior to death was 2.40±1.53 years. Most individuals (64%) had EOCD. AD (n=7) and FTLD (n=5) were the most common primary diagnoses, followed by DLB (n=3), VCI (n=2), corticobasal degeneration (n=1), PDD (n=1), autoimmune encephalitis (n=1), and epilepsy (n=1). Additionally, four individuals had co-pathologies These data highlight the clinical and pathological diversity of neurodegenerative disorders associated with mortality in this cohort.

**Figure 6.**
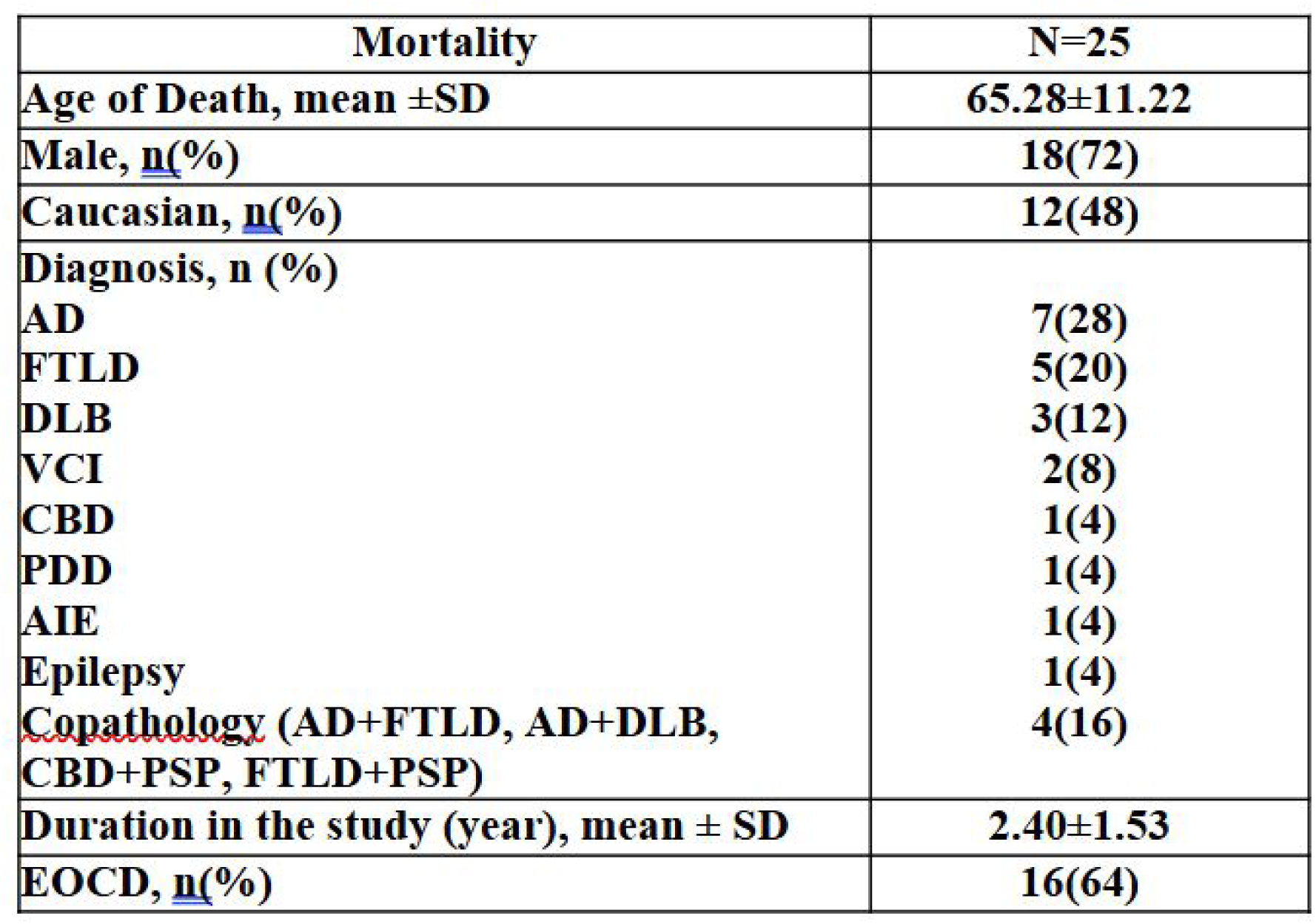
Mortality. Abbreviations: AD = Alzheimer’s disease; FTLD = frontotemporal lobar degeneration; VCI = vascular cognitive impairment; DLB = dementia with Lewy bodies; CBD = corticobasal degeneration: PDD= Parkinson’s Disease Dementia; AIE= Autoimmune Encephalitis; PSP= Progressive Supranuclear Palsy.

## Discussion

### Overview

As defined as the onset of dementia symptoms before the age of 65 years, young-onset dementia accounts for less than 10 % of all dementia. However, the disease burden has experienced an upward trend as global age standardized prevalence and incidence of young onset dementia increased from 93.39 and 16.24 per 100,000 persons in 1990 to 96.09 and 17.16 per 100,000 persons in 2021 (He et al., 2025). As most previous studies were focused on the late-onset dementia, developing a clinical cohort of younger individuals with cognitive impairment provides a comprehensive overview of their demographic, clinical, and biomarker profiles. Memory services/clinics vary massively in UK. This study also gives an example of memory services.

The CogNID Study aims to explore and better understand the role of cognitive and neuroimaging factors in the early diagnosis or neurodegenerative conditions. Our initial data shows the wide range of age and diagnoses which attend the memory clinics locally. The detailed analyses offer insights into the spectrum of cognitive impairment, the prevalence of co-existing medical conditions, and the diagnostic yield of neuroimaging and CSF biomarkers in a heterogeneous clinical population.

This cohort was predominantly comprised of individuals with early-onset cognitive symptoms (81.4% EOCD vs. 18.6% LOCD), with a mean age of 60.05 years. Both groups shared a similar male predominance and ethnic distribution. The percentage of Caucasian participants was lower than local census, given the exclusion criteria of ‘Not speaking English before the age of five years’ before last year. Cultural differences may influence help-seeking behaviour (Perry-Young et al., 2018), potentially resulting in an underrepresentation of certain groups within our sample, and across our clinic patient lists. Our cohort includes approximately 8% participants from ethnic minority backgrounds, while 28% have not disclosed their ethnicity. This highlights the challenges in capturing a fully representative sample. Although we amended the protocol inclusion criteria to remove being able to speak English before the age of five years, this was done at the end of the recruitment window included in this paper. Therefore, it is likely that we missed the opportunity to recruit more participants from minority ethnic backgrounds, and we need to be mindful of this for future studies. Factors such as ethnicity, diversity, educational level, and illiteracy are important to consider. Evidence suggests that individuals from ethnic minority groups are less likely to attend clinics (Sung et al., 2024) and participate in research studies (Haley et al., 2017).

### Mental Health, Cardiovascular Risk, and Modifiable Factors in Dementia

Notably, the high prevalence of depression and anxiety in our cohort (29.6%) underscores the complex interplay between neuropsychiatric symptoms and systemic comorbidities in individuals with cognitive impairment. Depression and anxiety are recognised both as early manifestations of neurodegenerative disease and as independent risk factors for progression to dementia. Longitudinal studies have shown that depression in MCI is associated with distinct clinical trajectories, with higher conversion rates to AD and VCI (Donovan et al., 2015; Wang et al., 2020). Moreover, meta-analyses, such as Yang et al. (2023) using UK Biobank data, report that depression increases the risk of developing dementia by approximately 1.5-fold. Anxiety has also been independently linked to the later development of Alzheimer’s disease, suggesting that early neuropsychiatric screening may offer opportunities for earlier intervention (Mah et al., 2015).

Beyond mental health, cardiovascular risk factors were also prevalent, with hypertension affecting 15.9% and T2DM affecting 9.8% of participants. These findings align with epidemiological data from the Framingham Heart Study and others, which have demonstrated strong associations between midlife hypertension, diabetes, and later cognitive decline (Skoog et al., 1996; Cheng et al., 2012). T2DM is associated with increased risk of both vascular cognitive impairment and Alzheimer’s disease, with systematic reviews confirming the elevated prevalence of MCI among diabetic populations (Biessels et al., 2006; Cheng et al., 2022).

Lifestyle factors also play a key role. Smoking has been consistently identified as a modifiable risk factor for both the development and progression of dementia. In younger populations particularly, smoking may contribute to earlier onset through mechanisms such as vascular injury, oxidative stress, and neuroinflammation. Given the significantly higher prevalence of current smoking among early-onset dementia cases in our cohort (6.3% vs. 0% in late-onset), memory clinics and brain health services should actively promote smoking cessation as a primary prevention strategy (Livingston et al., 2020; UK Brain Health Clinic guidelines., 2021).

Overall, these findings reinforce the importance of recognising and addressing modifiable health behaviours, cardiovascular risks, and mental health symptoms as part of a comprehensive, multidisciplinary approach to dementia diagnosis and management.

### Neurocognitive and Biomarker Assessment

Cognitive evaluation using the ACE-III revealed that a substantial majority (72.2%) of patients scored below the established threshold. These deficits were similarly distributed between EOCD and LOCD groups, although these have not been age adjusted.

The higher utilisation of lumbar puncture in the EOCD group (37.5% vs. 23.8%, p = 0.03) suggests that clinicians may be more inclined to employ advanced diagnostic tool when faced with atypical or diagnostically challenging presentations in younger patients. The high prevalence of abnormal Aβ 1-42/1-40 ratio (49.5%) and tau (t-tau 30.7% and p-tau-181 44.2%) profiles reinforces the utility of CSF studies, particularly in cases where traditional imaging modalities may be inconclusive.

In addition, biomarker-based classification offers opportunities for more precise treatment selection and prognosis prediction. Stratifying patients based on their amyloid (A) and tau (T) status allows for a nuanced understanding of disease stage and progression. For instance, individuals classified as A+T−—indicating abnormal Aβ but normal tau—are often considered to be in a preclinical or early prodromal stage of Alzheimer’s disease. This group may remain clinically stable for a longer period compared to those with both amyloid and tau abnormalities (A+T+), who are more likely to experience faster cognitive decline and progression to dementia (Eriksson et al., 2023). Recent study of CSF biomarkers suggested that a model disease staging using combination of CSF biomarkers can be used to predict prognosis (Sakvado et al., 2024). Thus, the integration of CSF biomarker profiles into clinical practice not only enhances diagnostic accuracy but also supports a personalised medicine approach, guiding therapeutic decisions and informing patients about their likely disease course.

### Subgroup study: Alzheimer’s Disease and Functional Cognitive Disorder

In the subgroup analysis, AD patients were older and exhibited more pronounced cognitive deficits than FCD groups. Despite similar rates of brain image and CSF biomarker testing between the groups, the marked difference in CSF biomarker abnormality as well as mortality. A retrospective study has shown that FCD patients had lower age at present, higher MMSE scores and Geriatric depression scale compared with neurodegenerative disorders (Borelli et al., 2022). FCD has been reported as common diagnosis in memory clinic that a quarter of patients received diagnoses of FCD from memory clinics associated with psychogenic symptoms in systematic review (McWhirter et al., 2020).

### Clinical and Diagnostic Considerations

For all research concerning possible and confirmed AD and dementia diagnoses, we follow the NICE guidelines (NICE, 2018). The guidelines include cognitive, radiological and CSF assessments. However, as most participants in our cohort are younger, with atypical presentations rarely observed in older groups, there are currently no specific guidelines tailored to younger populations. Therefore, we require further research into the diagnosis of those with EOCD, to create specific guidelines we can be sure cover all required elements.

The integration of neuropsychological testing, advanced neuroimaging, and CSF biomarker analysis provides a multidimensional approach to the diagnosis of cognitive disorders, particularly in a population where early-onset presentations may pose unique diagnostic challenges. The increased diagnostic yield in EOCD cases using lumbar puncture may reflect not only the diagnostic complexity in this subgroup but also a broader differential diagnosis considered by clinicians in younger patients.

Interestingly, co-pathologies have been observed in our cohort, underscoring the role of multiple pathological processes in the clinical manifestation of cognitive symptoms (Jack et al., 2024). In the Brains for Dementia Research study, histological evidence revealed that many participants exhibited co-pathologies, such as amyloidosis alongside TDP-43 proteinopathy (Selvackadunco et al., 2019). Emerging plasma biomarkers—such as GFAP and αSyn-SAA— as well as neuroimaging techniques, including MRI and FDG-PET, may aid in detecting additional pathologies implicated in disease progression, such as vascular brain injury, neuroinflammation, and α-synucleinopathy (Jack Jr. et al., 2024). The presence of co-pathologies is often associated with more rapid cognitive decline and a poorer prognosis (Robinson et al., 2021).

Plasma biomarkers such as phosphorylated tau-217 (p-tau-217) have quickly moved into clinical practice and trial eligibility criteria in countries including the United States, where both immunoassay and mass-spectrometry methods report high diagnostic accuracy for brain amyloid and tau pathology (Ashton et al., 2024). In the United Kingdom, a large-scale clinical trial has been launched to evaluate the accuracy, feasibility, and scalability of plasma p-tau-217 and other plasma assays in diverse real-world settings.

### Limitations

The sample for this study has limited generalisability as all participants werederived from memory service all within the East Midlands and limited to those selected for referral to the tertiary neurology-lead clinic. This recruitment process relies on patients being diagnosed and accessing these services. In addition, we exclude patients with known cause of cognitive difficulties (e.g., following major stroke, traumatic brain injury, or substance misuse); this may limit the consideration of co-pathologies often associated with dementia. Reporting of smoking and alcohol use within the cohort was likely underreported, possibly due to clinicians not routinely asking about these factors or patients feeling uncomfortable disclosing such information (Bisschop et al., 2025). We know that excessive alcohol consumption or smoking increases the risk of dementia (Livingston et al., 2020), and therefore this study may indicate that clinicians need to amend how they explore these habits with patients.

### Conclusions

Longitudinal data are essential for providing deeper insights into the prevalence and characteristics of cognitive disorders within tertiary referral centres. Our centre serves a diverse population, offering valuable opportunities to implement and evaluate diagnostic, preventive, and therapeutic strategies over time. The CogNID study highlights the heterogeneity of cognitive presentations in a young-to-midlife population and underscores the importance of a multidisciplinary diagnostic approach, incorporating cognitive assessments, neuroimaging, and biomarker profiling.

Although recruitment limitations—such as regional sampling and referral bias—must be acknowledged, the growing size and diversity of our cohort strengthen its future research potential. The challenges we encountered, including underrepresentation of minority ethnic groups and underreporting of lifestyle factors, provide important lessons for future study design and recruitment strategies.

As this is an ongoing study, with the number of participants now exceeding 600, future follow-up data will allow us to investigate longitudinal outcomes, explore predictors of disease progression, and assess the impact of interventions. Ultimately, these efforts aim to improve early diagnosis, personalise treatment pathways, and contribute to the development of specific clinical guidelines for early-onset dementia.

## Scope Statement

Our manuscript investigates the clinical, cognitive, neuroimaging, and biomarker profiles of early- and late-onset cognitive decline within a deeply phenotyped cohort. The study offers novel insights into the heterogeneity of cognitive decline in aging populations, with particular emphasis on early-onset cases, which remain under-recognized and diagnostically challenging. These findings align closely with Frontiers in Psychiatry – Aging Psychiatry’s focus on neuropsychiatric aspects of aging, dementia, and translational approaches to cognitive disorders in older adults. Our work contributes to advancing understanding and clinical recognition of age-related and atypical cognitive syndromes, supporting the journal’s mission to bridge research and practice in psychiatric care for the aging population.

## Conflict of interest statement

### The authors declare a potential conflict of interest and state it below

The author(s) declared that they were an editorial board member of Frontiers, at the time of submission. This had no impact on the peer review process and the final decision

### Credit Author Statement

**Abigail Rebecca Lee:** Data curation, Methodology, Resources, Visualization, Writing – original draft, Writing – review & editing. **Akram A. Hosseini:** Conceptualization, Data curation, Funding acquisition, Methodology, Project administration, Supervision, Validation, Visualization, Writing – review & editing. **Beili Shao:** Data curation, Formal Analysis, Investigation, Methodology, Project administration, Resources, Validation, Visualization, Writing – original draft, Writing – review & editing. **Bruno Gran:** Data curation, Methodology, Resources, Supervision, Writing – review & editing. **Elizabeta Blagoja Mukaetova-Ladinska:** Investigation, Validation, Visualization, Writing – review & editing. Hannah Sargisson: Data curation, Methodology, Writing – review & editing. **JeYoung Jung:** Data curation, Methodology, Writing – review & editing. Kehinde Junaid: Methodology, Writing – review & editing. **Permesh Dhillon:** Methodology, Writing – review & editing. **Peter Sellars:** Data curation, Methodology, Resources, Supervision, Writing – review & editing.

## Funding information

AAH has received research funding from the UK Medical Research Council (grant MR/T005580/1) through a Clinical Academic Partnership Award.

## Funding statement

The author(s) declare that financial support was received for the research and/or publication of this article.

## Ethics statements

### Studies involving animal subjects

Generated Statement: No animal studies are presented in this manuscript.

### Studies involving human subjects

Generated Statement: The studies involving humans were approved by This study has been reviewed and approved by the NHS East Midlands Derby Research Ethics Committee (18/EM/0292; IRAS: 250525). The studies were conducted in accordance with the local legislation and institutional requirements. The participants provided their written informed consent to participate in this study.

### Inclusion of identifiable human data

Generated Statement: No potentially identifiable images or data are presented in this study.

### Data availability statement

Generated Statement: The raw data supporting the conclusions of this article will be made available by the authors, without undue reservation.

### Generative AI disclosure

No Generative AI was used in the preparation of this manuscript.

## Conflict of Interest

None of the authors have conflict of interest.

## Author Contributions

**Akram A. Hosseini:** Conceptualization, Methodology, Validation, Writing – review & editing, Visualization, Supervision, Project administration, Funding acquisition. **Beili Shao:** Methodology, Data curation, Validation, Formal analysis, Investigation, Resources, Writing – original draft, Writing – review & editing, Visualization. **Abigail R. Lee:** Methodology, Data curation, Resources, Writing – original draft, Writing – review & editing, Visualization. **Permesh Dhillon:** Methodology, Writing – review & editing. **Kehinde Junaid:** Methodology, Writing – review & editing. **Bruno Gran:** Methodology, Data curation, Resources, Supervision, Writing – review & editing. **Peter Sellars:** Methodology, Data curation, Resources, Supervision. **Hannah Sargisson:** Methodology, Data curation, Writing – review & editing. **JeYoung Jung:** Methodology, Data curation, Writing – review & editing. **Elizabeta B. Mukaetova-Ladinska:** Writing – review & editing, Visualization, Investigation, Validation.

## Funding

AAH has received research funding from the UK Medical Research Council (grant MR/T005580/1) through a Clinical Academic Partnership Award; the National Institute on Aging, National Institutes of Health, USA (grant 1R56AG074467-01); and a research grant from Eisai, administered through her institution. AAH has received funding for Alzheimer’s education, training and advice from Biogen, Eisai and Lilly. JJ was supported by the Academy of Medical Sciences Springboard (SBF007\100077). BG receives personal compensation for consultancy from Merck, Roche, Biogen, Teva UK, GW Pharma, and Gilead Sciences. BG receives unrestricted research grants from Biogen Idec, Merck, Bayer Healthcare, Teva UK, Novartis, and Genzyme. BG receives support for the attendance of clinical and research conferences from Biogen, Merck, Bayer Healthcare, Teva UK, Novartis, Genzyme, and CelGene.

## Acknowledgments

The authors would like to thank all the participants and their families who have, and continue to, support this study.

## Notes

### Competing Interest Statement

The authors have declared no competing interest.

### Funding Statement

AAH has received research funding from the UK Medical Research Council (grant 611 MR/T005580/1) through a Clinical Academic Partnership Award; the National Institute on 612 Aging, National Institutes of Health, USA (grant 1R56AG074467‐01); and a research grant 613 from Eisai, administered through her institution.

